# How ‘Sturgeon’ guides the surgeon

**DOI:** 10.1101/2025.09.25.25336329

**Authors:** Mariska Sie, Oscar H.J. Eelkman Rooda, Mariëtte E.G. Kranendonk, Pieter Wesseling, Marc Pagès-Gallego, Lennart Kester, Marc van Tuil, Eric Strengman, Janneke Slijkoort-Blom, Eugène T.P. Verwiel, Arie Maat, Jasper van der Lugt, Jeroen de Ridder, Bastiaan B.J. Tops, Carlo Vermeulen, Eelco W. Hoving

## Abstract

For pediatric patients with central nervous system (CNS) tumors, primary treatment often involves neurosurgical tumor resection. Accurate diagnosis is crucial to perform the best-suited extent of resection. Since two years, Sturgeon, an AI-based validated intraoperative nanopore sequencing tool, has been fully integrated with frozen section analyses as standard of care in our nationwide centralized pediatric oncology hospital. This care evaluation is the first to demonstrate the impact of Sturgeon on neurosurgical decision-making. Sturgeon delivered correct diagnoses in 82 out of 94 consecutive patients (87.2%, <90 minutes), no diagnosis in 11.7% and 1 incorrect diagnosis. The diagnosis obtained by Sturgeon supported the intended surgical strategy (85.7%) or changed the strategy (14.3%) toward a more aggressive or limited resection. This resulted in only 3.2% second-look surgeries. In conclusion, intraoperative use of Sturgeon provides essential guidance toward the most optimal neurosurgical strategy and thereby has great potential to contribute to better clinical outcome.

## Introduction

Central nervous system (CNS) tumors are the most common solid tumors in children and still represent the leading cause of cancer-related death in the pediatric population^1^. Primary treatment often involves tumor resection in which the neurosurgeon needs to find the optimal balance between maximizing the extent of resection and minimizing the risk of neurological deficits^2^. The appropriate extent of resection depends on the tumor type, the availability of other effective treatment modalities, and the associated overall survival. For some tumor types, such as posterior fossa group A (PFA) ependymoma and atypical teratoid/rhabdoid tumor (AT/RT), complete neurosurgical resection (gross total resection, GTR) is crucial for improving oncological outcome and survival, underscoring the importance of optimal initial surgery to prevent the need for a second-look surgery^3–8^.In contrast, GTR is not necessarily the best surgical strategy for *e*.*g*., specific histomolecularly defined medulloblastomas^9^. Thus, an accurate diagnosis—whether preoperative or intraoperative—is critical for tailoring the surgical approach to each pediatric CNS tumor patient.

Currently, more than 100 pediatric CNS tumor (sub)entities are recognized^10–12^, but despite advances in medical imaging technology, an unequivocal diagnosis can often not be based on MRI imaging alone^13^. Consequently, tumor resection surgery is in various cases initiated without a clear diagnosis. Intraoperative frozen section analysis provides a provisional diagnosis, but cannot cover the complete gamut of CNS tumor types. Therefore, the diagnosis of the most fine grained (sub)entities are increasingly based on molecular characteristics of the tumor^14,15^, that cannot be detected based on hematoxylin-and-eosin (H&E) stained sections^16,17^. Two years ago, we launched Sturgeon, a deep-learned CNS tumor classification algorithm. By analyzing sparse methylation profiles derived from shallow nanopore DNA sequencing, Sturgeon is able to deliver an accurate methylation-based diagnosis during surgery^18^. We demonstrated the practical feasibility and reliability of intraoperative application of Sturgeon for pediatric and adult CNS tumors in different hospitals^18^. In the last two years, several other groups have also reported strategies for intraoperative nanopore sequencing diagnostics^19–23^, indicating that this approach is having a fundamental impact on the field^24–26^, but clear evidence of the clinical impact of intraoperative molecular classification is still lacking.

In the Princess Máxima Center for Pediatric Oncology, the national center for pediatric (neuro-)oncology in the Netherlands, Sturgeon has been validated and subsequently implemented in the routine intraoperative diagnostic workflow for well over two years. In this workflow, the tumor tissue obtained by the neurosurgeon is analyzed by H&E stained frozen sections in parallel with nanopore sequencing and subsequent classified using Sturgeon. The pathologist informs the neurosurgeon intraoperatively with a frozen section diagnosis followed by Sturgeon’s diagnosis, interpreted as an integrated histomolecular diagnosis.

To evaluate the impact of intraoperative sequencing on real-time neurosurgical decision-making, we performed a care evaluation of 94 consecutive pediatric patients that underwent a CNS tumor resection in the past two years. Sturgeon corroborated or refined the frozen section diagnosis, clarified unclear diagnoses or even prevented misdiagnoses. In 85.7% of cases in which Sturgeon generated a confident diagnosis, it supported the intended surgical strategy, while in 14.3% Sturgeon significantly changed the surgical strategy (14.3%). In these last cases, we provide representative examples to illustrate how Sturgeon guided the neurosurgeon toward a more aggressive or conservative surgical strategy. As each CNS tumor resection is unique, these descriptions are presented explicitly at the individual patient level. This demonstrates the nuances of Sturgeon’s impact during complex pediatric CNS tumor resection.

## Results

### Patient and tumor characteristics

In total, we performed concurrent intraoperative frozen section analysis and nanopore sequencing during tumor resection in 94 consecutive pediatric patients with primary CNS tumors in the Princess Máxima Center for Pediatric Oncology in the Netherlands. In these cases, tumor resection was multidisciplinary indicated. Patients with a known histomolecular diagnosis from a diagnostic biopsy or other previous surgeries were excluded. In a subgroup (the most recent 73 included patients), the impact of our diagnostic intraoperative workflow (Fig. 1) on the neurosurgical strategy was systematically evaluated.

**Fig. 1.**
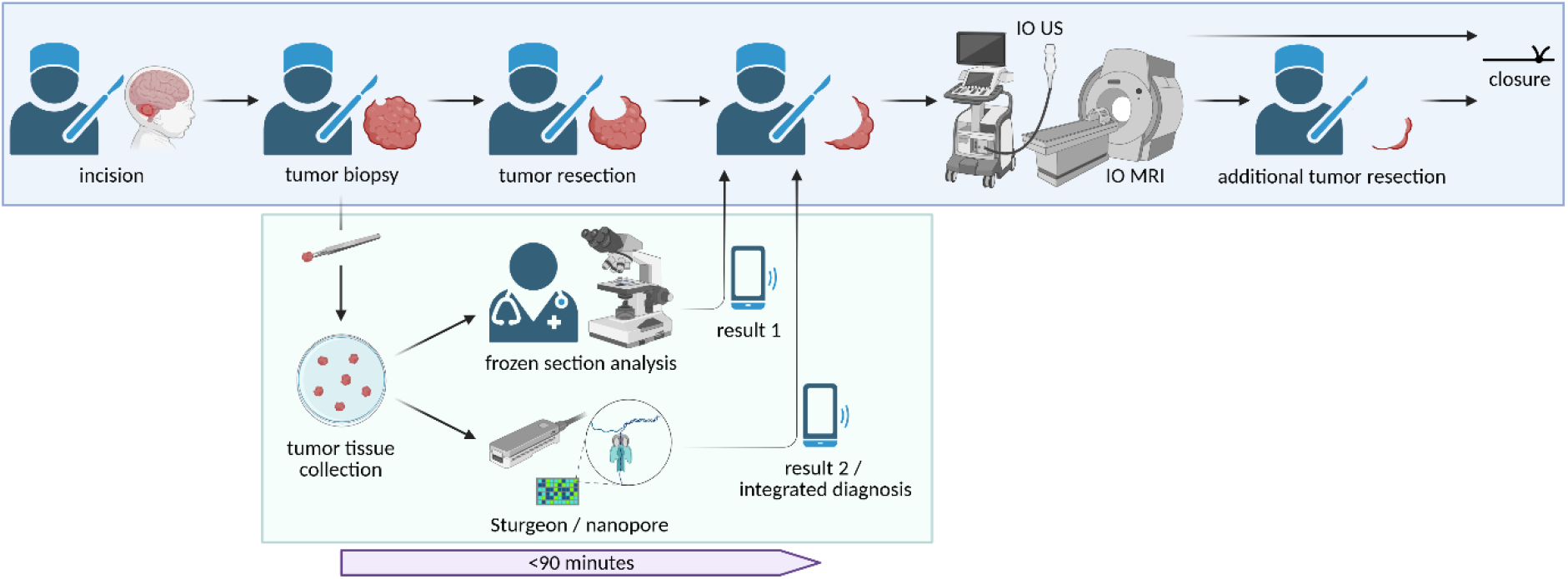
Intraoperative workflow. Intraoperative workflow combining frozen section analysis with Sturgeon’s nanopore sequencing. An integrated histomolecular diagnosis is provided within 90 minutes from the moment the tumor tissue was collected in the operating room until the results were communicated by the pathologist with the neurosurgeon(s) to inform real-time surgical decision-making. Intraoperative ultrasound (IO US) and intraoperative magnetic resonance imaging (IO MRI) were used to achieve the most optimal surgical strategy based on multiple factors including the provided intraoperative diagnosis, tumor location/invasiveness and patient characteristics.

Median age at surgery was 7.9 years (range 0.2 – 18.7) with a male predominance of 60.6%. The tumor characteristics are summarized in Figure 2, revealing a diverse patient group that reflects the heterogeneous pediatric CNS tumor population. In 89 patients (94.7%) the tumor was intracranially located and 5 patients (5.3%) had a spinal tumor. The most frequent postoperative diagnosis (conform the 5^th^ edition of the WHO Classification of Tumours of the Central Nervous System (WHO CNS5)^12^ using state-of-the-art diagnostics) was pilocytic astrocytoma (n = 22), followed by medulloblastoma (n = 19), ependymal tumor (n = 16) and high-grade glioma (n = 12).

**Fig. 2.**
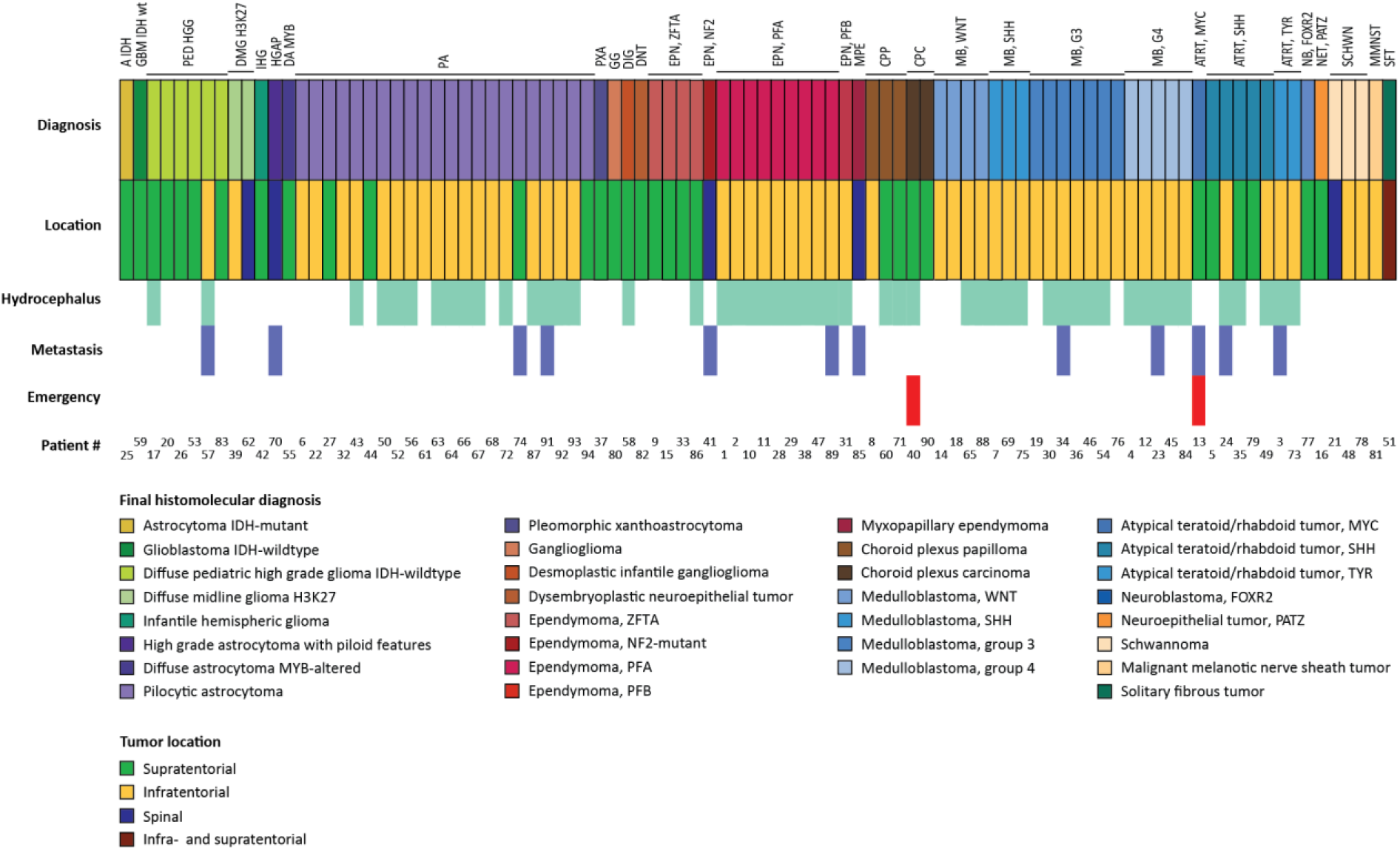
Tumor characteristics. Overview of the broad spectrum of final histopathological diagnoses (based on WHO CNS5 gold-standard diagnostics) in the total patient group (n = 94). This reflects the heterogeneity of the pediatric CNS tumor population. Craniopharyngioma and germ cell tumors were excluded. More than half of the patients (53.2%) were diagnosed with secondary hydrocephalus at presentation based on clinical symptoms and radiological findings. Different treatment strategies were performed in 42 out of 50 patients with hydrocephalus (84.0%): external drainage (57.1%), endoscopic third ventriculostomy (4.8%), dexamethasone 4-6 mg/m2/day (35.7%) and 1 patient (2.4%) received a ventriculoperitoneal shunt abroad which was removed during tumor resection surgeryat our hospital. Metastatic disease at diagnosis was observed on craniospinal MRI in 12 out of 94 patients (12.8%). Tumor resection itself was consideredas an emergency surgery (< 24 hours after radiological diagnosis) in 2 patients (2.1%) due to tumor mass effect with acute clinical deterioration.

### Intraoperative equipment

For the most optimal surgical performance, we used intraoperative ultrasound (IO US, BK medical) and Brainlab neuronavigation, respectively in 88.3% and 71.3% of all 94 cases, mainly combined as navigated ultrasound. Intraoperative 3T MRI (IO MRI) was used in 59 of the surgeries (62.8%), and in 12 of those patients (20.3%) IO MRI prompted additional tumor resection because of suspected residual tumor (pilocytic astrocytoma n = 7; high-grade glioma n = 2; glioneuronal tumor n = 1; AT/RT n = 2). Intraoperative neuromonitoring was used in 16 patients (17.0%) with either an infratentorial (n = 11) or spinal tumor (n = 5).

### Preoperative and intraoperative diagnostics

To evaluate the performance of intraoperative sequencing, we compared preoperative radiology, intraoperative frozen section analysis and Sturgeon-based diagnoses. During the intraoperative diagnostic workflow, the frozen section diagnosis was communicated as a first result followed by Sturgeon’s result interpreted in the context of the microscopic features observed in the frozen section and communicated with the neurosurgeon(s) as a second, more integrated intraoperative diagnosis. When the frozen section diagnosis and the Sturgeon classification were in disagreement, the frozen section was carefully re-evaluated, which usually prompted a revision of diagnosis communicated by the pathologist on the condition that it would be appropriate for clinical and (revised) radiological features. This demonstrates the complementary value of both diagnostic tools and emphasizes the importance of a well-integrated histomolecular workflow.

We found that in 54 of the 94 patients (57.4%), the most likely diagnosis formulated by experienced neuroradiologists was in concordance with the final integrated CNS5 WHO histomolecular diagnosis (Fig. 3). In another 6 patients (6.4%) the most probable diagnosis based on radiology overlapped with the final diagnosis but lacked specificity. In 34 patients (36.2%) the most likely diagnosis based on MRI was incorrect. Intraoperative fresh frozen section analyses were performed by one or two experienced (neuro)pathologists and were consistent with the final postoperative diagnosis in 66 patients (70.2%). A more general diagnosis was given in 14 cases (14.9%) which overlapped with the final diagnosis, but lacked the more fine grained subclass designation, for example ‘embryonal’ in case of medulloblastoma or AT/RT and ‘glioma’ without further differentiating between low-grade or high-grade. The frozen section diagnosis was incorrect in 14 cases (14.9%).

**Fig. 3.**
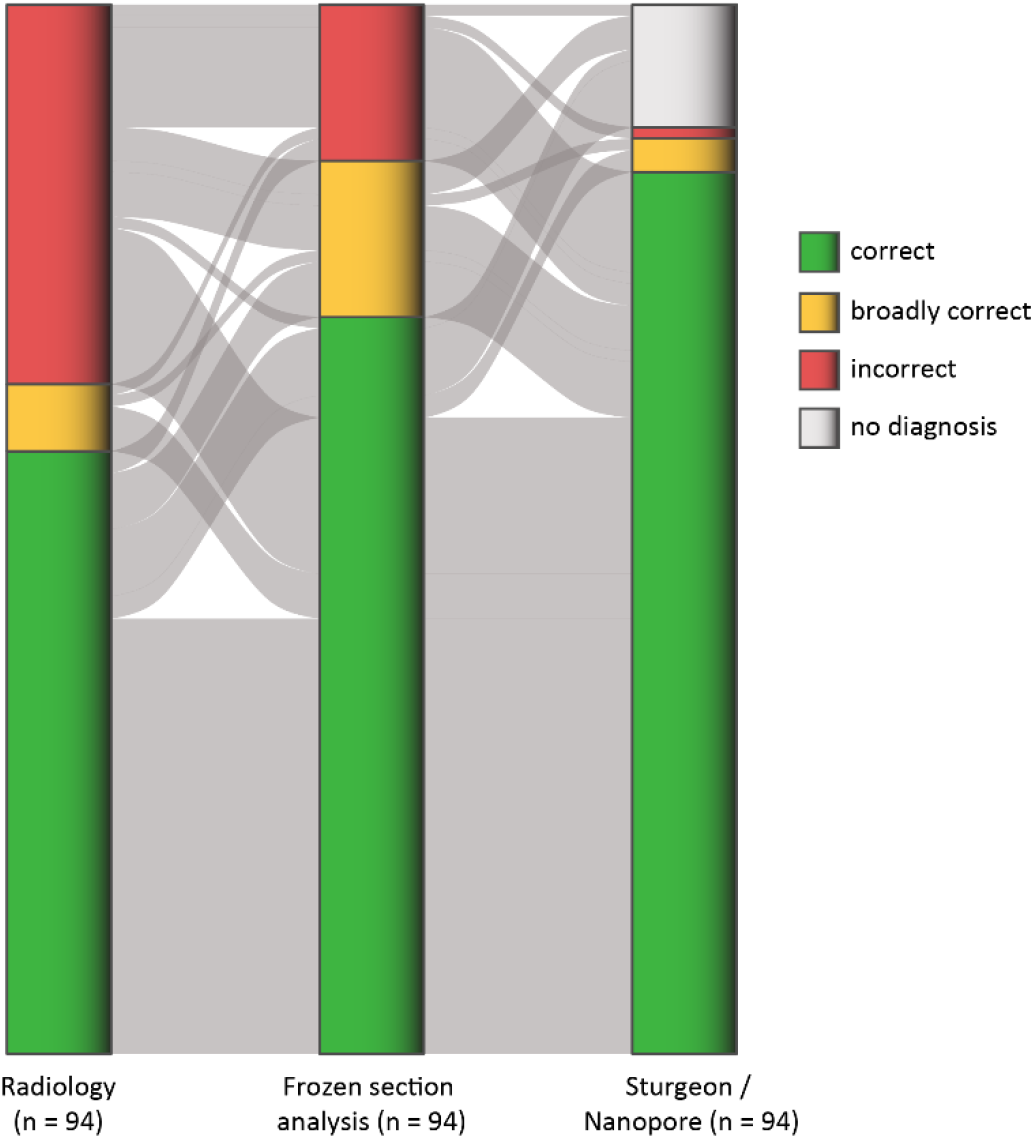
Preoperative and intraoperative diagnostics. Alluvial plot showing the results of the different diagnostic tools over time for the total patient group (n = 94). In 54 out of 94 (57.4%) cases the most likely radiological diagnosis was consistent with the postoperative final CNS5 WHO histomolecular diagnosis, compared with 66 out of 94 (70.2%) correct intraoperative frozen section diagnoses and 82 out of 94 (87.2%) correct diagnoses including subclasses by intraoperative Sturgeon’s nanopore sequencing. All 82 nanopore sequencing results reached a confidence score of ≥0.90 (and ≥0.95 confidence score in 77 out of 82 cases). ‘Broadly correct’ radiological and frozen section diagnoses overlapped with the final histomolecular diagnosis in 6.4% (6/94) and 14.9% (14/94) of cases respectively, but lacked specificity. In 3 out of 94 cases (3.2%) Sturgeon provided the correct tumor type (family) but an incorrect subclass: PFB ependymoma instead of PFA, AT/RT-TYR instead of AT/RT-SHH and medulloblastoma group 4 instead of group 3.

In contrast, Sturgeon provided a correct diagnosis including subclassification in 82 out of 94 patients (87.2%) within 90 minutes after the first tumor tissue was collected in the operating room (Fig. 3). A confidence score ≥0.95 was reached in the majority of these cases. In 5 out of 82 cases (6.1%) Sturgeon reached a confidence score between 0.90 and 0.95, but in those cases results were also relayed to the surgeon after careful consideration alongside the radiological, surgical and frozen section findings. In 3 cases, the subclass designation provided by Sturgeon was incorrect, specifically a PFB ependymoma instead of PFA, AT/RT TYR instead of AT/RT SHH and medulloblastoma group 4 instead of group 3. Except for this incorrect medulloblastoma subgroup classification, all other 18 medulloblastoma cases, including their subgroups, were diagnosed correctly (94.7%).

Sturgeon did not reach a confident diagnosis in 11 out of 94 cases (11.7%), which mostly included pilocytic astrocytoma or other low-grade gliomas or glioneuronal tumors. Low confidence classifications likely occurred due to a low tumor fraction. One case was diagnosed with an entity that is not present in the current Sturgeon classifier (neuroepithelial tumor with *MN1::PATZ* fusion^27^). Lastly, an incorrect diagnosis was suggested by Sturgeon in only 1 out of 94 cases (1.1%). This concerned a patient with a hypervascular tumor centered around the tentorium with mass effect on both left cerebral and cerebellar hemisphere. The radiological and frozen section analyses pointed in the direction of meningioma. The Sturgeon classification alternated between meningioma and solitary fibrous tumor (SFT), settling on meningioma. This was inconsistent with the definitive diagnosis, which was SFT. The misdiagnosis was possibly due to substantial presence of meningeal tissue in the sample that was used for Sturgeon analysis. This can be prevented by taking tumor tissue biopsies more centrally avoiding the superficial parts invading surrounding tissue like meninges.

### Sturgeon’s impact on neurosurgical strategy

For a subgroup (n = 73) of the most recent cases we systematically analyzed the direct consequence of Sturgeon’s intraoperative diagnosis on the neurosurgical strategy. We observed that in all cases where Sturgeon generated an intraoperative diagnosis (63/73 cases, 86.3%) the neurosurgical strategy was influenced. In 54 out of 63 cases (85.7%), Sturgeon supported the surgical strategy and even changed the surgical strategy in 9 out of 63 cases (14.3%).

### Sturgeon’s impact on pediatric brain tumor surgery – 3 illustrative cases

Intraoperative Sturgeon diagnosis resulted in a subtle neurosurgical strategy change in a young patient with a large posterior fossa tumor (5.9 × 3.7 × 3.6 cm) arising from the vermis (Fig. 4, patient 24). The radiological differential diagnosis included AT/RT, embryonal tumor with multilayered rosettes (ETMR) and ‘atypical medulloblastoma’. AT/RT requires the most extended surgical resection to improve survival^5^. For medulloblastoma, on the other hand, there is no prognostic benefit to gross total resection (GTR) over near total resection (NTR) when the likelihood of neurological morbidity is high^9^. ETMR is relatively rare and although GTR is associated with a longer overall survival compared with subtotal resection (STR), the number of cases is too small to distinguish if GTR would be beneficial over NTR^28^. Frozen section analysis indicated an embryonal tumor without further differentiation, which encompasses all radiological diagnoses (AT/RT, ETMR and medulloblastoma). Interestingly, Sturgeon was decisive, settling on AT/RT SHH (consistent with the final diagnosis) and guided toward a more aggressive approach. In this patient, intraoperative (IO) MRI showed residual tumor which was additionally resected, achieving a complete surgical resection, MR0 (no visible tumor on MRI), and preventing a second-look surgery.

**Fig. 4.**
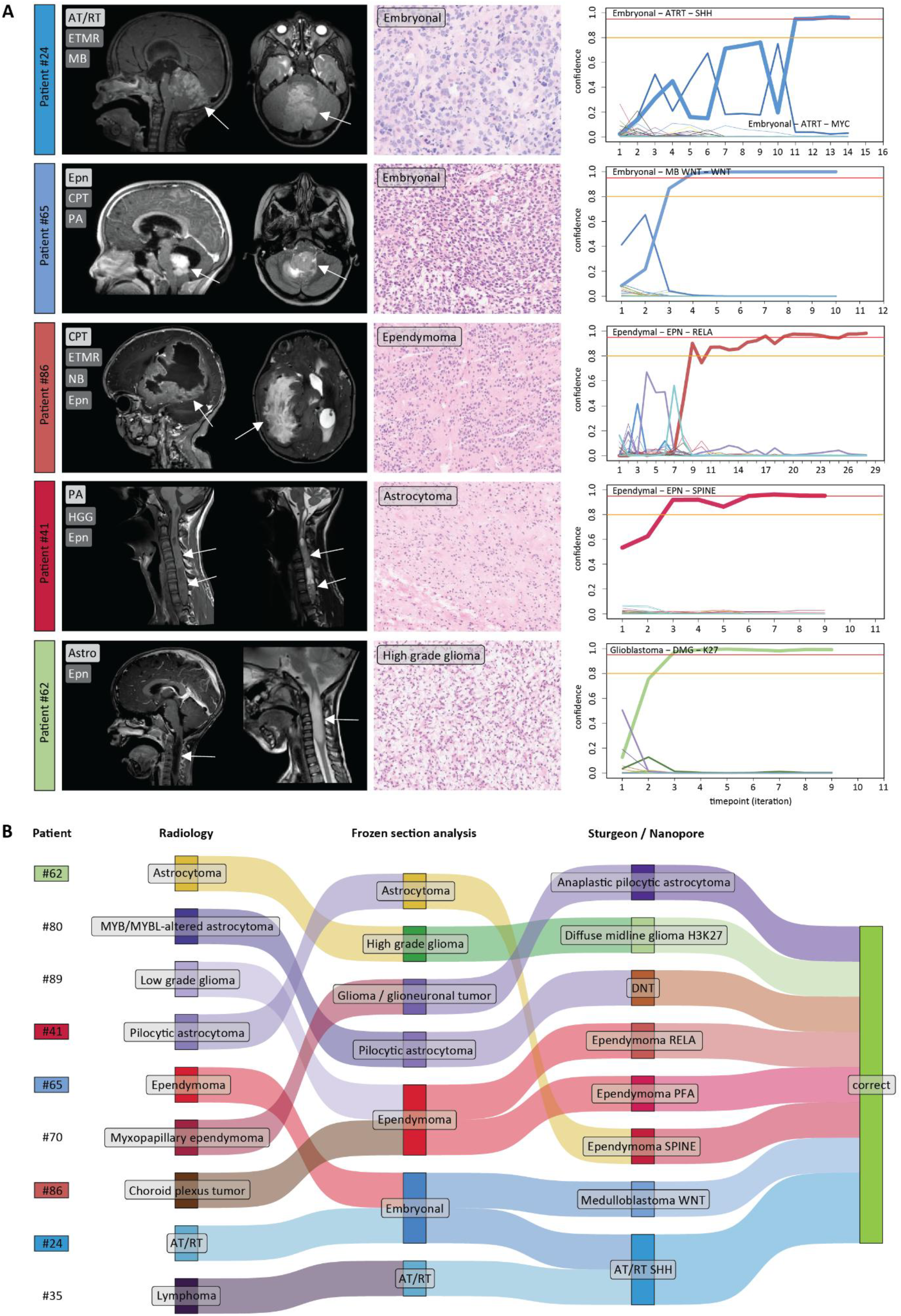
Sturgeon’s impact on neurosurgical strategy. In our subgroup analysis (n = 73), in all 63 out of 73 cases (86.3%) where Sturgeon generated an intraoperative diagnosis by nanopore sequencing, this influenced the neurosurgical strategy. In 54 out of 63 cases (85.7%) Sturgeon supported the intended surgical strategy and in 9 out of 63 cases (14.3%) it changed the strategy toward a more aggressive or limited resection. **A**, Three illustrative intracranial cases and two spinal cases are shown with preoperative magnetic resonance T1 (with or without contrast) and T2 images (intracranially sagittal and axial images; for spinal cases both sagittal images). Arrows demark tumors. Corresponding intraoperative frozen section H&E staining analyses resulted in broad diagnoses while Sturgeon was decisive or even overruled the frozen section diagnosis guiding toward the most optimal surgical strategy if possible. **B**, Sankey diagram showing the most likely diagnosis per modality forthe 9 patients where an adjusted surgical strategy was indicated by intraoperative diagnostics. Patient numbers are shown on the left. Columns indicate from left to right the potential diagnosis based on radiology, frozen section histology and Sturgeon classification. The final column indicates whether the Sturgeon classification was postoperatively confirmed with the final CNS5 WHO histomolecular diagnosis. MB denotes medulloblastoma; Epn, ependymoma; CPT, choroid plexus tumor; PA, pilocytic astrocytoma; NB, neuroblastoma; HGG, high-grade glioma; Astro, astrocytoma.

In contrast, Sturgeon can also lead to a more conservative surgical approach. This was the case in a patient with a tumor in the fourth ventricle, radiologically diagnosed as ependymoma, choroid plexus tumor or, less likely, pilocytic astrocytoma (Fig. 4, patient 65). For all these tumor entities GTR is indicated with the nuance that, for ependymoma and choroid plexus carcinoma, GTR significantly improves overall survival when followed by adjuvant treatment modalities^3,4,6,7,29–31^. For choroid plexus papilloma and pilocytic astrocytoma, GTR alone is often curative ^32–36^. However, for pilocytic astrocytoma, STR can still lead to a long-term survival^36^. In contrast to radiology, frozen section analysis showed features of an embryonal tumor and Sturgeon confirmed a WNT-activated medulloblastoma, which was in line with the definitive postoperative diagnosis. For this reason, based on Sturgeon’s result, a more conservative approach was performed at the level of the cerebellar peduncles where the tumor invaded (SR1: suspected residue/possible local invasion; but not visible on MRI: MR0)^9^.

Importantly, despite the availability of accurate intraoperative diagnoses the optimal surgical strategy for the specific tumor entity could not always be achieved, due to tumor location and invasiveness. For example, in a young patient with a large hemispheric tumor located in the right lateral ventricle and extending in the quadrigeminal plate cisterns. The radiological differential diagnosis included choroid plexus carcinoma/papilloma, embryonal tumor and, with a lower likelihood, ependymoma (Fig. 4, patient 86). Frozen section analysis and Sturgeon diagnoses were in agreement, suggesting ependymoma and *ZFTA* fusion ependymoma, respectively. Based on this intraoperative diagnosis^6^, an aggressive surgical approach was performed. However, due to attachment of the tumor to the central cerebral veins, combined with significant intraoperative blood loss (3200 cc) and multiple transfusions, STR (SR2, MR2) was the maximum achievable extent of resection. This highlights the necessity of a tailored surgical approach, where patient-specific factors can dictate the final extent of resection despite an intraoperatively informed surgical strategy.

### Sturgeon’s impact on pediatric spinal tumor surgery – 2 illustrative cases

Sturgeon also had a major impact on the surgery of spinal cases. The first patient presented with an extensive atypical intramedullary tumor with both cystic and solid components at the level of C3 until Th4 (Fig. 4, patient 41). The radiological differential diagnosis included pilocytic astrocytoma, high-grade glioma and ependymoma. Although frozen section analysis suggested astrocytoma, Sturgeon contradicted this diagnosis with a high confidence score for spinal ependymoma. Sturgeon’s diagnosis was more in line with the intraoperative neurosurgical findings of a better defined plane between tumor and displaced spinal cord. The diagnosis spinal ependymoma was postoperatively confirmed (ependymoma with *NF2* mutation, CNS WHO grade 2). In order to maximize the chance of overall survival, based on this diagnosis^7,37,38^, a more radical resection was performed than preoperatively anticipated (SR0 and MR0 instead of SR1).

In contrast, a more conservative approach was indicated in the second patient with an intramedullary tumor, this time reaching from the level of C6 to just above the foramen magnum. Based on radiology this tumor was preoperatively diagnosed as a glial tumor, astrocytoma more likely than ependymoma (Fig. 4, patient 62). Frozen section analysis resulted in high-grade glioma and Sturgeon confirmed and further refined the diagnosis toward a diffuse midline glioma (DMG) H3K27. Due to this DMG diagnosis, which are incurable^39^, it was essential to limit the intervention, and only perform an open biopsy (SR3 and MR3). Sturgeon’s diagnosis thus prevented any further risk for possible neurological deficits.

### Intraoperative complications and second-look surgery

In our patient population, with use of the intraoperative diagnostic workflow (which incorporates frozen section analysis, Sturgeon, intraoperative imaging and neuromonitoring), we observed less intraoperative complications compared with literature. Intraoperative complications such as cranial nerve injury occurred in 2 out of 94 patients (2.1%). Both patients with cranial nerve injury presented with a large, multicystic tumor in the posterior fossa (one PFA ependymoma and one AT/RT SHH) that showed lateral and ventral extension. In 1 out of 94 patients (1.1%), resection of a large choroid plexus carcinoma was complicated by unexpected excessive blood loss (4300 cc) with cardiovascular instability. Median intraoperative blood loss in the total group was reported to be 200 cc with a wide range of 0 – 4300 cc and in 31.9% of the cases intraoperative blood transfusion was given. Previous studies report a total intraoperative complication rate of 6.4% (10 out of 156 patients) and 6.7% (19 out of 284 patients), both in the context of pediatric CNS tumor surgery^40,41^.

Although the primary goal of intraoperative diagnosis is to improve safety and reduce morbidity, it may also contribute to reducing the need for second-look surgeries. Indeed, in only 3 out of 94 (3.2%) patients a second-look surgery within 30 days was performed, while in a retrospective study this was done in 12 out of 161 patients (7.5%)^42^. Two out of three of the performed second-look surgeries were two-staged tumor resection surgeries where a second surgery was preferred because of safety, rather than being indicated due to unexpected residual tumor tissue. In one patient the tumor was too extensive to be resected in a single surgical approach. In another case, a two-staged tumor resection was preferred after a blood loss of 1200 cc and multiple blood transfusions though without cardiovascular instability. Only 1 out of 94 cases (1.1%) presented with as an unexpected residual tumor, which was detected on postoperative MRI (IO MRI was not available during this specific surgery). All three additional surgeries were multidisciplinary indicated and performed within only one week after the first resection. Sturgeon contributed to this quick decision-making. The surgeries resulted in GTR and an uneventful recovery. Median time intervals reported in the literature for second-look surgery are 15 and 27.3 days, with transient morbidity^42,43^, which potentially leads to a delay in adjuvant therapy and worsening of long-term outcome.

### Hospital admission and communication

Pediatric CNS tumor patients require a multidisciplinary approach, so after performing the best surgical strategy, it is essential to schedule adjuvant treatment correctly without delay to maximize prognosis. Postoperative hospital admission is considered a useful measure of healthcare quality and reflects the postoperative course. In the total patient group (n = 94), we observed a median hospital stay after tumor resection surgery of only 6.5 days (range 2 – 239) compared with a mean of 8.7 days (12.5 SD) in high-case volume hospitals and 11.0 days (16.7 SD) in low-case volume hospitals^44^. In the Princess Máxima Center, the length of admission may be overestimated because adjuvant chemotherapy is sometimes initiated during the initial hospitalization, prolonging the hospital stay.

Secondly, the final histomolecular diagnosis must be discussed by a multidisciplinary team prior to informing the patient and their parents about additional therapies and follow-up. This is preferably done in a timely manner to limit the time spent in uncertainty. It also provides the ability to perform all necessary preparations for patients that require adjuvant therapy and prevents treatment delay. Remarkably, the median length between surgery and communication of the multidisciplinary tumor board results with patients and their parents was 8 days (range 0 – 17 days after surgery); 62 times (66.0%) during the initial admission, 32 times (34.0%) in the outpatient clinic. In cases with a correct intraoperative Sturgeon result (n = 82) versus those where Sturgeon failed to provide a diagnosis (n = 11), the histomolecular diagnosis including therapeutic plan were more frequently communicated during the initial hospital stay (70.7% versus 36.4%, Fisher’s exact test, p = 0.038) and often sooner after surgery (median 8 days, range 0 – 17 days versus median 13 days, range 2 – 15 days, Mann-Whitney U test, p = 0.018). The gold standard for a final histomolecular diagnosis in our center remains a combination of histopathological examination, immunohistochemistry (IHC), EPIC methylation array profiling, whole exome sequencing, and whole transcriptome sequencing^14^. However, when Sturgeon’s result was concordant with both frozen section analysis as well as clinical and radiological features, diagnostic confidence was considered high and earlier communication of a histomolecular diagnosis with patients and families became feasible. Nevertheless, assessment of confirmative and prognostically relevant molecular alterations remain part of the gold standard diagnostic workup.

## Discussion

This is the first clinical evaluation of Sturgeon, an intraoperative DNA methylation-aware nanopore sequencing classifier to support real-time neurosurgical decision-making in pediatric neuro-oncology. In pediatric CNS tumors, which are often deep-seated and difficult to distinguish preoperatively^13,16,17^, the surgical strategy must be tailored to the unique factors of each patient. We show that Sturgeon provides a valuable tool to enhance diagnostic confidence during crucial intraoperative moments, with the diagnosis being a key factor in guiding the surgical strategy.

For pediatric CNS tumors where GTR is associated with better overall survival, such as in AT/RT and ependymoma^3–7^, Sturgeon’s diagnosis supported a more aggressive surgical approach. Conversely, for tumors where a more conservative surgical strategy is clinically justified, like in WNT-activated medulloblastoma and DMG^9,39^, the diagnosis aided in confirming that approach. While the optimal strategy was not always possible due to factors like tumor location and invasiveness, Sturgeon contributed to informed decision-making in the majority of cases and actually changed the surgical strategy taken in close to 15% of the cases.

The successful integration of Sturgeon into our surgical workflow, alongside other intraoperative tools like neuronavigation, ultrasound, MRI and neuromonitoring and frozen section analysis, resulted in a reduced incidence of second-look surgeries compared with those documented in literature^42^. This low rate of unplanned second surgeries can contribute to better patient outcomes by reducing morbidity, avoiding delays in the start of adjuvant therapy, and potentially improving long-term prognosis. The use of Sturgeon also enabled earlier communication of a histomolecular diagnosis to the patients and families, reducing the time spent in uncertainty with an unclear prognosis. Taken together, the integration of Sturgeon is a crucial step in optimizing care for these vulnerable patients. A key strength of Sturgeon is its universality and low investment cost^18^. Sturgeon can be used without computationally expensive retraining and therefore runs on inexpensive consumer-grade hardware, making it a promising tool for implementation in hospitals with limited financial resources globally. This is especially important given the high prevalence, morbidity and mortality of pediatric CNS tumors worldwide^1,45,46^.

Sturgeon may benefit from further improvement to address the limited number of inconclusive or incorrect diagnoses by improving robustness to low tumor purity and increasing the number of tumor entities in the reference data. Future studies should focus on specific tumor types and use a multicenter approach to validate these findings to further advance the field of intraoperative molecular diagnostics.

## Methods

### Patients and evaluation

Since May 16, 2023, after internal validation of Sturgeon^18^, a dual diagnostic neuro-oncological workflow including intraoperative frozen section analyses and ultra-fast nanopore sequencing using Sturgeon has been implemented as standard of care in the Princess Máxima Center for Pediatric Oncology in the Netherlands (Fig. 1). This intraoperative diagnostic workflow is applied to all pediatric patients (< 19 years of age) with primary CNS tumors and the multidisciplinary indication for tumor resection surgery. Intraoperatively generated results are subsequently communicated by the (neuro)pathologist with the pediatric neurosurgeon(s) in the operating room enabling real-time decision-making during CNS tumor surgery.

Since January 1, 2024 we systematically reported the intraoperative impact of Sturgeon on real-time neurosurgical decision-making (n = 73) as follows: having no impact, being supportive in the surgical strategy or changed the surgical strategy. The neurosurgical strategy was partly evaluated by the intended extent of resection before and during surgery (before/after frozen section and nanopore results) according to the latest recommendation of the International Society of Pediatric Oncology (SIOP). This includes an integrated surgical and radiological grading system. Surgical impression on resection was therefore graded from SR0 to SR3: SR0 (complete resection), SR1 (rim like residual), SR2 (bulky residual), SR3 (biopsy)^47^. Radiological assessment was graded MR0 to MR3 based on MRI findings: MR0 (complete resection), MR1 (rim-like residual ≤ 3 mm), MR2 (residual > 3 mm in any section), MR3 (residual > 50% of initial volume) (Table 1)^47^.

**Table 1.** Extent of resection as evaluated intraoperatively and on postoperative contrast-enhanced MRI to be performed within 48h (max 72h) after surgery.

|  |  |
| --- | --- |
| SR 0 | Total resection, no residue |
| SR 1 | Suspected residue, possible local invasion |
| SR 2 | Solid residuum (to be defined by postoperative MRI) |
| SR 3 | Tumor volume unchanged, biopsy |
| MR 0 | No visible tumor |
| MR 1 | Rim enhancement or signal abnormality (matching the tumor) at the operation site only (“Rim”), $\leq 3$ mm in any of the dimensions and equivocal for tumor residue |
| MR 2 | Residual tumor measuring $> 3$ mm in all 3 dimensions (greater than MR1, less than MR3) |
| MR 3 | No significant change to preoperative tumor size (“minimal change”) |

We performed a care evaluation by analyzing all intraoperative generated data from consecutively treated patients between May 16, 2023 until May 1, 2025. Patients and/or their parents/guardians provided written informed consent for the biobank (reviewed and approved by the Medical Ethics Review Committee, see International Clinical Trials Registry Platform: NL7744) (n = 94). The intraoperative impact evaluation was performed on the last 73 included patients. All evaluations were approved by the Biobank and Data access committee (BDAC) of the Princess Máxima Center for Pediatric Oncology.

Patients with known histomolecular diagnosis derived from previous biopsy or (partial) tumor resection were excluded. Moreover, patients with radiological diagnosis of craniopharyngioma or germ cell tumors were excluded because craniopharyngioma can typically be histologically diagnosed without molecular analyses^12^ and germ cell tumors were not yet represented in the Sturgeon classifier used for this evaluation.

### Sample collection, registration and splitting

For intraoperative sample collection, we performed fresh tumor tissue biopsies (at least 5 × 5 × 5 mm tissue in total) in the beginning of the tumor resection, preferable not directly from the tumor rim. This material was collected fresh in the operating room and delivered on ice to the pathology department. In the meantime, the neurosurgeon(s) continued the surgery. Upon receipt of the intraoperative specimen, the pathologist macroscopically divides the tissue into two paired subsamples designated for frozen section analysis and nanopore sequencing, respectively. For each of two tissue fragments, one portion was processed for frozen section analysis, while the adjacent portion was simultaneously subjected to DNA extraction for nanopore library preparation. This parallel processing allows for efficient turnaround without delaying sequencing.

### Frozen section histology

Intraoperative pathological evaluation was performed using frozen section analysis. Fresh tissue specimens were promptly processed using the FlashFREEZE™ system (Miltenyi Biotec), which enables rapid and standardized cryopreservation. The tissue was embedded in MMC (methylcellulose-milk cryoprotectant) to ensure optimal preservation of tissue architecture, and snap-frozen within seconds to preserve morphological integrity. Frozen tissue blocks were sectioned at 3 µm using a cryostat and stained with hematoxylin and eosin (H&E) for immediate microscopic evaluation by an experienced pathologist. The frozen section diagnosis provided preliminary information on tumor type and malignancy grade which was communicated directly to the neurosurgical team to support intraoperative decision-making.

### Nanopore sequencing

The subsamples designated for nanopore sequencing (1 – 2 subsamples per patient) were processed using a previously described method to isolate DNA (Qiaamp Tissue DNA extraction kit)^18^. Once the frozen sections were prepared and reviewed microscopically, the pathologist evaluated tumor content and morphology to determine which subsample contained the highest tumor fraction. Based on this assessment, the corresponding adjacent fragment—already undergoing DNA isolation was selected for library prep and sequencing. Library prep was conducted with the LSK-RBK114.24 library prep kit (Oxford nanopore technologies) and sequenced on a MinION flowcell.

### Intraoperative classification

Sequencing data was collected using a dedicated GridION device (Oxford Nanopore Technologies). Minknow software was configured to sequence with basecalling disabled. A Docker container with the required software was run in parallel to sequencing. This Docker runs Guppy basecalling, Modkit-based methylation-state extraction and the Sturgeon classifier whenever a new pod5 file was written to disk. Sturgeon’s results were communicated directly to the pathologist as soon as preliminary classification became available and were made definitive when it reached a confidence score ≥0.95^18^. These molecular findings were interpreted in the context of the histomorphological features observed in the frozen section analysis and communicated by the pathologist with the neurosurgeon(s) to inform real-time surgical decision-making. Also the already communicated frozen section results were revised in light of the Sturgeon diagnosis. The revised diagnosis was considered the integrated intraoperative diagnosis. In case of permanent discrepancy, Sturgeon’s results were followed if it could be in line with tumor location, re-evaluation of preoperative MRI and intraoperative surgical findings.

### Reproduced timelines

To retrospectively compare the timing of intraoperative sequence experiments, we developed a script that repeated the analysis performed intraoperatively from raw data (pod5) files one-by-one in chronological order, using the file time-stamps as a timepoints. In cases where the timestamps were overwritten, the times were not added to the plot.

### Validation of Sturgeon classification

The accuracy of the nanopore/Sturgeon classification was assessed by comparison with the final integrated WHO CNS5 diagnosis. This reference diagnosis was established through a comprehensive histomolecular workup, incorporating histology, immunohistochemistry (IHC), EPIC methylation array profiling, whole exome sequencing, and whole transcriptome sequencing^14^.

## Data Availability

All data produced in the present work are available upon reasonable request to the authors

## Author contributions

M.P.-G., J.d.R. and C.V. designed and created the Sturgeon algorithm. M.S., O.H.J.E.R., M.E.G.K., P.W., B.B.J.T., C.V. and E.W.H. developed the intraoperative workflow. M.S., O.H.J.E.R. and E.W.H. performed tumor resection surgeries, provided samples and translated the intraoperative diagnosis toward the most optimal surgical strategy. J.v.d.L. provided clinical input. M.v.T., E.S., J.S.-B., E.T.P.V., A.M., L.K., and C.V. collected and prepared intraoperative samples for intraoperative frozen section histology and performed intraoperative nanopore sequencing. M.E.G.K. and P.W. performed intraoperative frozen section analysis and provided the intraoperative and final integrated diagnosis. M.S., M.E.G.K., and C.V. collected and interpreted the data. M.S., O.H.J.E.R., M.E.G.K., C.V. and E.W.H. wrote the manuscript with input and critical review of all other authors.

