## Supplementary figure 1 for "How ‘Sturgeon’ guides the surgeon"

### Supplementary figures

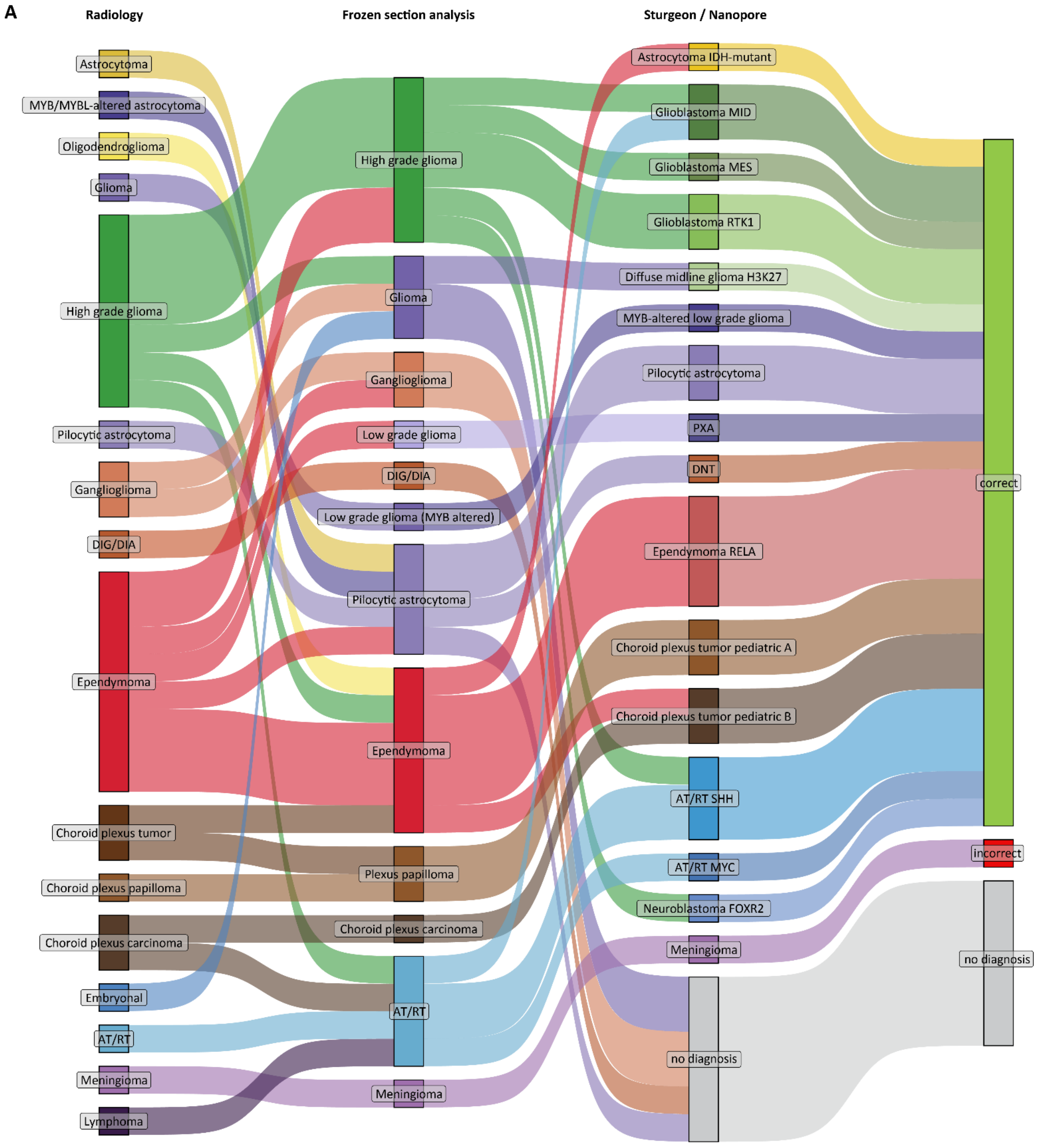

B

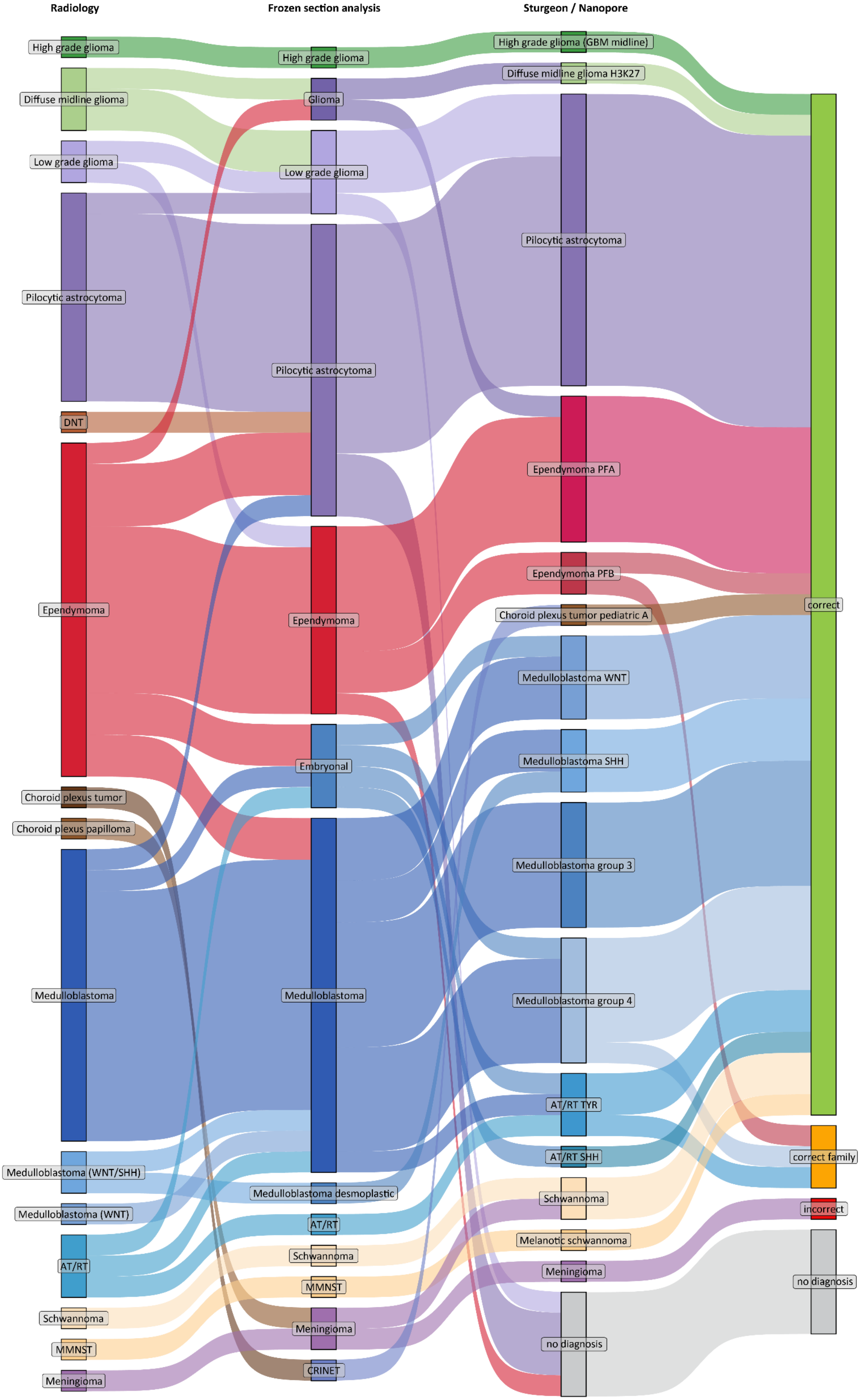
Radiology
Frozen section analysis
Sturgeon / Nanopore
High grade glioma
Diffuse midline glioma
Low grade glioma
Pilocytic astrocytoma
DNT
Ependymoma
Choroid plexus tumor
Choroid plexus papilloma
Medulloblastoma
Medulloblastoma (WNT/SHH)
Medulloblastoma (WNT)
AT/RT
Schwannoma
MMNST
Meningioma
High grade glioma
Glioma
Low grade glioma
Pilocytic astrocytoma
Ependymoma
Embryonal
Medulloblastoma
Medulloblastoma desmoplastic
AT/RT
Schwannoma
MMNST
Meningioma
CRINET
High grade glioma (GBM midline)
Diffuse midline glioma H3K27
Pilocytic astrocytoma
Ependymoma PFA
Ependymoma PFB
Choroid plexus tumor pediatric A
Medulloblastoma WNT
Medulloblastoma SHH
Medulloblastoma group 3
Medulloblastoma group 4
AT/RT TYR
AT/RT SHH
Schwannoma
Melanotic schwannoma
Meningioma
no diagnosis
correct
correct family
incorrect
no diagnosis

C

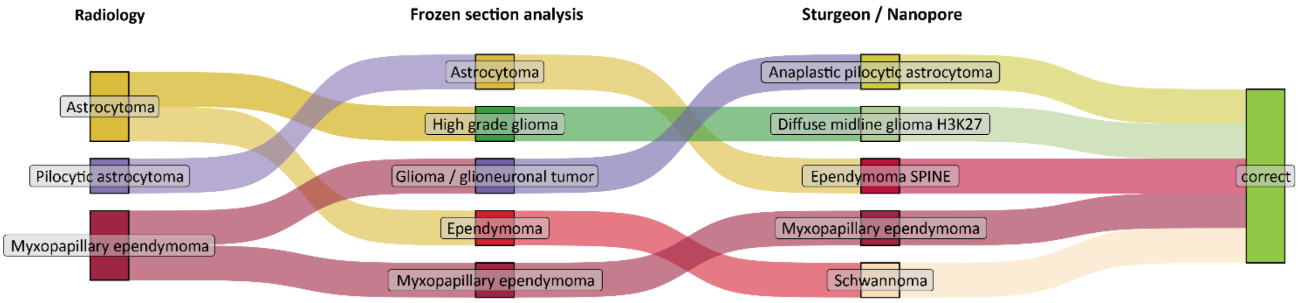


**Fig. S1** | Sankey diagrams of patients with (**A**) supratentorial located tumors (n = 32), (**B**) infratentorial (n = 58) and (**C**) spinal tumors (n = 5). Columns indicate from left to right the most likely preoperative radiological diagnosis based on MRI, intraoperative frozen section diagnosis and Sturgeon's intraoperative result. The final column indicates whether the Sturgeon classification was postoperatively confirmed by final CNS5 WHO histomolecular diagnosis. In three 'correct family' cases the tumors were correctly diagnosed as ependymoma, medulloblastoma and AT/RT however with an incorrect subclass: PFB ependymoma instead of PFA, medulloblastoma group 4 instead of group 3 and AT/RT TYR instead of AT/RT SHH. Note one patient out of 94 cases was incorrectly diagnoses by Sturgeon as meningioma instead of solitary fibrous tumor and is shown twice as the tumor was located at the level of the tentorium with supratentorial and infratentorial tumor mass.
